# Progressively Widening Healthcare Costs in Long COVID Over Five Years

**DOI:** 10.64898/2026.02.24.26346985

**Authors:** Jingya Cheng, Alaleh Azhir, Jiazi Tian, Jeffrey G. Klann, Shawn N. Murphy, Hossein Estiri

**Affiliations:** Department of Medicine, Massachusetts General Hospital, Boston, MA, USA; Department of Medicine, Brigham and Women’s Hospital, Boston, MA, USA

## Abstract

**Background:** Long COVID affects millions worldwide, yet the long-term trajectory of healthcare costs remains poorly characterized. Prior studies with limited follow-up have documented elevated but stable excess costs, leaving uncertainty about whether the economic burden attenuates or persists over time.

**Methods:** We conducted a retrospective cohort study using electronic health record data from 12 hospitals and 20 community health centers (January 2018 through December 2024). Adults with documented SARS-CoV-2 infection were classified as having Long COVID using a validated precision phenotyping algorithm or as controls without Long COVID. We used two-part generalized estimating equation models to estimate adjusted quarterly healthcare costs over 20 quarters, decomposed costs into visit frequency and cost-per-visit components, and conducted subgroup and sensitivity analyses accounting for differential mortality.

**Results:** Among 143,544 adults (27,986 with Long COVID; 115,558 controls), the adjusted excess quarterly cost for Long COVID widened progressively rather than attenuating, increasing from $79 (95% CI, $48–$118) at baseline to $236 (95% CI, $176–$287) at quarter 19 – a threefold increase in the cost differential. Long COVID was associated with 20% higher odds of any healthcare utilization (OR, 1.20; 95% CI, 1.18–1.23) and 30% higher costs when care was accessed (cost ratio, 1.30; 95% CI, 1.25–1.35). Visit frequency diverged over time, reaching 44% higher utilization by quarter 19, while cost-per-visit premiums remained stable. Excess costs concentrated in the upper distribution tail (99th percentile difference: $8,482). The widening trajectory was consistent across subgroups defined by hospitalization status, sex, and comorbidity burden. Cumulative 5-year excess costs were $7,124 per Long COVID patient after mortality adjustment.

**Conclusions:** Contrary to assumptions of post-acute recovery, Long COVID is associated with progressively widening healthcare costs over five years, driven primarily by increasing utilization rather than care intensity, suggesting an evolving chronic disease burden with substantial and growing economic implications.

## Introduction

Post-acute sequelae of SARS-CoV-2 infection (PASC), commonly referred to as Long COVID, is a multisystem disorder characterized by persistent or new symptoms that continue weeks to months after the initial infection.^1,2^ The condition affects an estimated 400 million people globally, with annual economic costs approaching $1 trillion, approximately 1% of the global economy.^3^ In the United States, projections incorporating lost productivity and quality of life reach $3.7 trillion.^4^ Direct healthcare costs represent a substantial component of this burden, with empirical studies reporting excess annual expenditures ranging from $176 to $2,683 per patient depending on population characteristics, healthcare settings, and measurement approaches.^3–7^

Despite growing attention to the economic impact of Long COVID, critical gaps limit our understanding. Most studies have followed patients for only 6 to 12 months,^7–9^ Most studies have followed patients for only 6 to 12 months, documenting elevated but stable excess costs that have been interpreted as reflecting a post-acute recovery phase, with the implicit assumption that the economic burden would attenuate as patients recover or adapt to chronic symptoms. Whether this assumption holds over longer follow-up remains unknown. Additionally, methodological heterogeneity, including variation in Long COVID definitions, cost components captured, and analytic approaches, hinders evidence synthesis.^7^ The dedicated ICD-10-CM code for post-COVID conditions demonstrates poor sensitivity, potentially leading to substantial underestimation when used for case identification.^10,11^

We conducted a retrospective cohort study of 143,544 adults with documented SARS-CoV-2 infection, using electronic health records data from 10 hospitals and 20 community health centers in New England, to examine healthcare costs associated with Long COVID over a five-year period. Using a validated phenotyping algorithm that prioritizes causal attribution through systematic exclusion of pre-existing conditions,^12^ we identified patients with Long COVID and compared their cost trajectories with those of infected patients who did not develop the condition. We employed two-part longitudinal models with pre-pandemic baseline data to quantify excess costs, characterize temporal patterns in cost accumulation, decompose costs into utilization and intensity components, and assess variation across demographic and clinical subgroups. This study provides the longest follow-up of Long COVID healthcare costs to date and tests whether excess costs attenuate, stabilize, or widen over time, a distinction with direct implications for health system planning and resource allocation..

### Background

Understanding the healthcare cost burden associated with long COVID is essential for health system planning, resource allocation, and policy development. A growing body of literature has examined healthcare costs attributable to long COVID and post-COVID conditions across diverse populations, healthcare settings, and geographic regions (**Table 1**). These studies vary substantially in their methodological approaches, including differences in PASC definitions, follow-up durations, cost measurement methods, and comparison groups. Early economic estimates suggested individual annual healthcare costs of approximately $9,000 (range: $3,700–$14,000) based on treatment cost parallels with myalgic encephalomyelitis/chronic fatigue syndrome (ME/CFS), contributing to an estimated $3.7 trillion total economic impact in the United States alone.^4^

**Table 1.** Healthcare Cost Studies in Long COVID and Post-COVID Conditions.

Published studies examining healthcare costs associated with Long COVID or post-COVID conditions.
| Study | Population | Setting/Country | Follow-up | PASC Definition | Excess/Annual Cost | Key Findings |
| --- | --- | --- | --- | --- | --- | --- |
| Cutler (2022)<sup>4</sup> | Estimated US Long COVID population | National economic estimate, USA | Annual | Chronic symptoms consistent with ME/CFS | \$9,000/year | Based on ME/CFS treatment cost parallels, part of \$3.7 trillion total US economic impact estimate |
| Koumpias et al. (2022)<sup>7</sup> | 250,514 COVID-19 patients | US commercial, public, and uninsured | 6 months | Post-acute sequelae $\geq 6$ months | \$223.60/month (\$2,683/year) | Costs are highest in ages 45-64 (\$73.43/month) and $\geq 65$ (\$60.49/month); persistent through the entire period |
| Pike et al. (2023)<sup>8</sup> | 197,412 adult cases; 11,636 child cases | US commercial insurance | 6 months | PCC; new problems $\geq 4$ weeks post-infection | \$393/month (Adults)<br>\$208/month (Children) | Among adults, excess medical costs during 1, 3, and 6 months were \$393, \$930, and \$1,562. Among children, excess medical costs during 1, 3, and 6 months were \$208, \$549, and \$1,011, respectively. |
| Chambers et al. (2023)<sup>9</sup> | 17,178 insured members | US health plans | 12 months | COVID-19 diagnosis | \$41.61 per member per month (~\$219/year) commercial | Costs elevated through 12 months; outpatient primary driver; propensity score matched |
| Sagy et al. (2023)<sup>13</sup> | ~640,000 case-control pairs | Integrated HMO, Israel | 15 months | Validated computable phenotype | \$7.7/month (7.6% increase); \$5.0/month for non-hospitalized | DiD methodology; monthly net excess cost; \$92.40/year |
| Yang et al. (2025)<sup>16</sup> | 30,122 adults with Long COVID | Primary care, France | Mean 13 months | WHO definition: symptoms $\geq 3$ months | €163/year excess (~\$176 USD); €2,443 total/year | 97.3% had GP visits; 62.4% nursing care; the highest costs were in the first year post-diagnosis |
| Sander et al. (2025)<sup>17</sup> | 159,817 persons with COVID-19 | Population-based, Ontario, Canada | 360 days | PCC; symptoms >12 weeks | \$2,553 CAD (~\$1,900 USD/year) | Phase-based analysis; 77% acute costs inpatient; 42% postacute costs inpatient; survival-adjusted |
| Lin et al. (2024)<sup>18</sup> | 52,988 Long COVID | UK OpenSAFELY primary care EHR | 2 years | Coded diagnosis or symptom clusters | £1,035/year excess (~\$1,300 USD) | 49% more healthcare utilization; £2,562/year total vs. £1,527 for controls |
| Łukomska et al. (2025)<sup>15</sup> | 41 studies synthesized | Multiple countries | Variable (6-24 months) | Variable definitions | 7.6-13.1% monthly cost increase (DiD); 1.3-4.0x higher vs. controls | Costs persist 15+ months; inpatient cases are 2-10x higher burden than non-hospitalized |
| Al-Aly et al. (2024)<sup>3</sup> | ~400 million globally | Worldwide | Ongoing | Long-term health effects of SARS-CoV-2 | ~\$1 trillion annually (global) | ~1% of global economy; includes direct and indirect costs; lost productivity major component |

Empirical studies using claims data and electronic health records have quantified excess healthcare costs ranging from modest increases of $7.70 per patient per month in integrated healthcare systems^13^ to substantially higher estimates exceeding $2,500 annually in populations with clinically diagnosed long COVID.^14^ A systematic literature review synthesizing 41 studies found that individuals with long COVID had significantly elevated healthcare resource utilization and costs compared to controls without a COVID-19 diagnosis, with a 7.6–13.1% increase in total healthcare costs per person per month as assessed by difference-in-differences analysis.^15^ Healthcare costs have been shown to persist for 15 or more months following infection, with hospitalized patients experiencing 2–10 times higher cost burden than those managed in outpatient settings.^7,9,15,16^

The heterogeneity in cost estimates across studies reflects several important methodological considerations in real-world evidence generation for long COVID, such as variations in case definitions (ranging from symptom-based criteria to ICD-10 coded diagnoses), differences in healthcare system structures and cost accounting methods, selection bias in study populations, and challenges in distinguishing long COVID-attributable costs from pre-existing conditions and general post-infection healthcare seeking behavior. Despite these limitations, the consistent finding of elevated healthcare utilization and costs across multiple healthcare systems and countries underscores the substantial economic burden of long COVID and highlights the urgent need for preventive strategies, effective treatments, and appropriate allocation of healthcare resources.^4,15^

## METHODS

### Study Design and Data Source

We conducted a retrospective cohort study using electronic health record data from the Mass General Brigham (MGB) health system, spanning from January 1, 2018, to December 31, 2024. The study cohort was derived from the validated PASC cohort (P2RC), which comprises integrated longitudinal clinical and demographic data from over 195,000 patients across 10 hospitals and 20 community health centers in Massachusetts. We employed a longitudinal cohort design to compare healthcare costs between patients with and without Long COVID following SARS-CoV-2 infection. The study incorporated a pre-pandemic baseline period (January 1, 2018, to December 31, 2019) to control for pre-existing healthcare utilization patterns and an analysis period (January 1, 202,0 to December 31, 2024) to capture post-infection costs across 20 consecutive quarters.

### Study Population

We included all adult patients (≥18 years) with documented SARS-CoV-2 infection, confirmed by either positive polymerase chain reaction or antigen testing, or by ICD-10 diagnosis codes for COVID-19. Patients satisfied a minimum data completeness score^19^ to ensure that they receive most of their care from the integrated healthcare system. Patients were stratified into two groups based on the development of post-acute sequelae of SARS-CoV-2 (PASC), commonly referred to as Long COVID. To ensure adequate data for longitudinal modeling and baseline covariate assessment, we required all patients to have: (1) at least one healthcare encounter during the baseline period; (2) at least one healthcare encounter during the analysis period; and (3) a minimum of two total encounters across all time periods.

PASC was identified using a validated phenotyping algorithm consistent with definitions from the World Health Organization and the U.S. National Academies of Sciences, Engineering, and Medicine. PASC was defined as an infection-associated chronic condition that persisted for more than two months after the acute infection episode and was not explained by alternative diagnoses. The phenotyping algorithm incorporated ICD-10 diagnosis codes, clinical symptoms, and temporal patterns. We applied an exclusion-based approach: a symptom was classified as PASC only if it emerged following the index infection, and no prior clinical diagnosis documented in the electronic health record from 2017 onward could account for it. For example, new-onset dyspnea was classified as PASC only in the absence of preexisting asthma or chronic obstructive pulmonary disease. Additional clinical expertise-based filters were applied to enhance the specificity of the PASC phenotype. Patients who developed PASC within 12 months of their index SARS-CoV-2 infection comprised the PASC group, while those who did not develop PASC within this timeframe served as the comparison group.

Given the cohort was derived from the P2RC, a validated PASC cohort identified using a precision phenotyping algorithm, there were no missing data for the primary outcome or covariates.

### Primary Outcome

The primary outcome was quarterly healthcare costs, defined as the sum of all healthcare expenditures per patient per calendar quarter from January 1, 2020, through December 31, 2024. Healthcare costs were derived from institutional billing records and included total payments from all payer sources: hospital payments, ambulatory surgery center payments, Medicare payments, and patient out-of-pocket costs.

### Cost Estimation

Healthcare costs were estimated using standardized national payment rates from the Centers for Medicare and Medicaid Services (CMS). We obtained procedure-level payment data from the CMS Hospital Outpatient Prospective Payment System (OPPS) annual policy files, which provide national average total payment amounts established by Medicare for healthcare services. For each encounter in the electronic health record, we extracted all associated procedure codes recorded as Current Procedural Terminology (CPT) or Healthcare Common Procedure Coding System (HCPCS) codes. These codes were then mapped to the corresponding national average total payment amounts from the CMS Procedure Price Lookup tables.

Encounter-level costs were calculated by summing the Medicare payment amounts for all procedures performed during that encounter. For encounters classified as inpatient admissions, we applied payment rates from the Inpatient Prospective Payment System; for outpatient encounters, we applied rates from the OPPS. When multiple procedure codes were documented for a single encounter, we aggregated all associated payments to derive the total encounter cost. Patient-quarter costs were then calculated by summing all encounter-level costs with service dates falling within each calendar quarter.

This approach provides standardized cost estimates that reflect resource utilization intensity based on procedures performed, independent of variation in negotiated payment rates across payers, geographic regions, or institutions. By using Medicare national average payments as a common metric, we reduce confounding from differential reimbursement patterns between groups and enhance comparability with other studies using similar standardization approaches. However, this method captures only procedure-based costs and does not include professional fees for evaluation and management services, laboratory tests, diagnostic imaging interpretation, or outpatient pharmacy expenditures. Additionally, Medicare payment rates may not reflect true economic costs or payments from commercial insurers, which are typically higher. All cost estimates were adjusted for inflation to 2024 U.S. dollars using the medical care component of the Consumer Price Index.

For each patient-quarter observation, we calculated total healthcare costs by summing all encounters and services with start dates falling within that quarter. Patient-quarters with no recorded healthcare utilization were assigned zero cost.

To address extreme outliers while preserving clinically meaningful high-cost cases, we applied winsorization at the 99.5th percentile of positive costs within the analysis cohort. Calendar quarters without any documented healthcare encounters were assigned a cost of zero. For the primary analysis, costs were censored at the time of death to isolate expenditures incurred during life.

### Covariates

Covariates were selected a priori based on established associations with healthcare utilization and costs. These included age at the index SARS-CoV-2 infection date (mean-centered with a quadratic term to capture nonlinear effects), sex, race (categorized as White, Black, Asian, or Other), ethnicity (Hispanic vs. non-Hispanic), the Charlson Comorbidity Index (mean-centered), and baseline quarterly healthcare costs. To control for pre-existing healthcare costs associated with underlying health status, we calculated baseline healthcare utilization during the pre-pandemic period. Baseline costs were defined as the mean quarterly cost across all encounters occurring between January 1, 2018, and December 31, 2019. They were log-transformed after adding one dollar to account for zero values, which addressed the skewed distribution of the baseline costs.

### Statistical Analysis

The primary analysis implemented a two-part regression model using a generalized estimating equations (GEE) framework to account for within-patient correlation in repeated quarterly cost measurements. The two-part model included two components: (1) the probability of incurring any healthcare costs and (2) the magnitude of costs conditional on having any utilization.

In the first part, we used logistic regression within a GEE framework with an exchangeable working correlation structure to model the probability of any healthcare use (cost >$0) in a given quarter by *logit*(*P*(*Cost* > 0)) = *β*_0_ + *β*_1_(*LC*) + *β*_2_(*Quarter*) + *β*_3_(*LC* × *Quarter*) + ***X**γ*. In the second part, among quarters with positive costs, we used gamma regression with a log link, also within a GEE framework with an exchangeable working correlation structure *log*(*E*[*Cost* | Cost > 0]) = *α*_0_ + *α*_1_(LC) + *α*_2_(*Quarter*) + *α*_3_(*LC* × *Quarter*) + ***X**δ*, to model the magnitude of costs. ***X*** represents a vector of covariates including age (centered at cohort mean, with quadratic term to capture non-linear age effects), sex, race (White, Black, Asian, Other), ethnicity (Hispanic, Non-Hispanic), Charlson Comorbidity Index (centered), hospitalization status during acute infection, and log-transformed baseline quarterly healthcare costs (to control for pre-pandemic utilization patterns). The PASC-by-time interaction allowed the cost trajectory to differ between groups over the follow-up period.

For each quarter, we calculated adjusted predicted mean costs for each group by averaging individual-level predictions across all patients. We then computed the difference in adjusted predicted costs between the PASC and non-PASC groups, along with 95% confidence intervals derived using robust standard errors.

### Cost Distribution Analysis

To characterize heterogeneity in cost impacts across the expenditure distribution, we constructed a patient-quarter–level panel in which each COVID-positive individual contributed up to 20 quarters of follow-up (0–19 quarters after January 2020). Quarterly costs were aggregated at the patient-quarter level, and censored at death using individual-specific death□quarter indices. To describe the distributional differences in healthcare utilization, we computed the empirical 25th, 50th (median), 75th, 90th, 95th, and 99th percentiles, as well as the mean of quarterly spending for LC and No LC groups for individuals with expenditures. We calculated the absolute difference and percentage change between Long COVID group and patients without Long COVID at each percentile to identify whether excess costs were uniformly distributed or concentrated among high-cost patients.

### Visit Frequency

To distinguish whether increased costs were driven by higher healthcare utilization (more frequent encounters) or greater intensity of care per encounter (higher costs when care was accessed), we implemented a decomposition framework using two additional generalized estimating equation models. First, We modeled the number of unique healthcare encounters per quarter using Poisson regression GEE with log link and exchangeable correlation structure, adjusting for the same covariate set used in the primary analysis. Second, among patient-quarters with at least one visit, we fit a Gamma GEE model with a log link to model cost per visit. We generated predicted trajectories for visit counts and cost per visit using the approach described for the primary cost predictions, including bootstrap confidence intervals.

Quarters were defined relative to January 1, 2020, and visit counts were obtained as the number of unique encounters per patient-quarter.

### Subgroup Analysis

To assess the generalizability of Long COVID’s cost trajectory across patient populations, we conducted stratified analyses by infection severity (hospitalized vs. non-hospitalized during acute COVID-19), sex (male vs. female), and comorbidity burden (Charlson Comorbidity Index <3 vs. ≥3). For each subgroup, we generated predicted quarterly costs using the two-part GEE models in the primary analysis, holding other covariates at reference values (age centered at mean, female sex for non-sex stratifications, White race, non-Hispanic ethnicity, non-hospitalized status for non-severity stratifications, and mean baseline cost). We calculated the excess cost for each quarter and subgroup, with 95% confidence intervals derived via parametric bootstrap with 400 replications drawing from the joint distribution of model parameters. The primary objective was to evaluate the consistency of the temporal cost differences pattern across clinically relevant subgroups rather than to test for statistically significant differences between subgroups. We present excess cost trajectories by subgroups to show the effect across populations.

### Sensitivity Analysis

To assess robustness with respect to differential mortality between groups, we conducted a sensitivity analysis incorporating survival probabilities into population-level expected costs. We estimated survival functions S(t) stratified by PASC status using the Kaplan-Meier method, with follow-up beginning January 1, 2020. For each quarter t, we calculated population-level expected costs as the weighted sum of two components: (1) mean costs among survivors at quarter t, weighted by S(t), and (2) mean end-of-life costs among decedents, weighted by the probability of death in quarter t. End-of-life costs were defined as total costs incurred in the final two quarters before death. This approach accounts for the fact that patients who die incur substantial terminal costs and are subsequently censored from the survivor cost estimates.

### Patient and Public Involvement

Patients and the public were not involved in the design, conduct, reporting, or dissemination plans of this research.

### Sample Size and Power

With more than 140,000 patients contributing approximately 3 million observations, we had greater than 99% power to detect cost ratios as small as 1.10 between groups at a two-sided alpha level of 0.05, assuming a coefficient of variation of 0.5 for healthcare costs.

All statistical tests were two-sided, and P values less than 0.05 were considered to indicate statistical significance. Analyses were performed using R software, version 4.3.1 (R Foundation for Statistical Computing).

## Results

A total of 143,544 adults met the inclusion criteria, including 27,986 individuals with Long COVID and 115,558 comparators without Long COVID. Table 2 summarizes demographic and clinical characteristics during the 2018–2019 baseline period. The mean age of the cohort was 53.6 years. Overall, 62.8% of participants were female; 77.0% were White, 8.1% were Black, and 5.1% were Hispanic. Among patients with PASC, the mean age was 56.1 years, and 64.0% were female. The racial and ethnic composition was as follows: 76.8% were White, 8.8% were Black, and 4.9% were Hispanic. The mean Charlson Comorbidity Index was 2.42 in the overall cohort and 3.14 among those with PASC.

**Table 2.** Summary statistics of the study population.

|  | Long COVID<br>(N=27986) | No Long<br>COVID<br>(N=115558) | P-value<br>(LC vs. NoLC) | Overall<br>(N=143544) |
| --- | --- | --- | --- | --- |
| **Age, years** Mean (SD) | 56.2 (17.0) | 53.0 (16.8) | <0.001 | 53.6 (16.9) |
| **Sex** |  |  |  |  |
| Female | 17905 (64.0%) | 72264 (62.5%) | <0.001 | 90169 (62.8%) |
| Male | 10081 (36.0%) | 43294 (37.5%) | <0.001 | 53375 (37.2%) |
| **Race** |  |  |  |  |
| Asian | 899 (3.2%) | 4437 (3.8%) | <0.001 | 5336 (3.7%) |
| Black | 2453 (8.8%) | 9203 (8.0%) | <0.001 | 11656 (8.1%) |
| Other | 3150 (11.3%) | 12839 (11.1%) | 0.495 | 15989 (11.1%) |
| White | 21484 (76.8%) | 89079 (77.1%) | 0.258 | 110563 (77.0%) |
| **Ethnicity** |  |  |  |  |
| Hispanic | 1368 (4.9%) | 5989 (5.2%) | 0.047 | 7357 (5.1%) |
| Non-Hispanic | 26618 (95.1%) | 109569 (94.8%) | 0.047 | 136187 (94.9%) |
| **Charlson Comorbidity Index Mean (SD)** | 3.14 (2.78) | 2.25 (2.34) | <0.001 | 2.42 (2.46) |
| **Mortality** | 1503 (5.4%) | 2873 (2.5%) | <0.001 | 4376 (3.0%) |
| **Baseline median quarterly cost, \$ Mean (SD)** | 1,030 (2,390) | 1010 (2450) | <0.001 | 1020 (2440) |
| **Baseline mean visits per quarter (2018-2019) Mean (SD)** |  |  |  |  |
| Encounters with at least a procedure code | 1.58 (1.25) | 1.51 (1.17) | <0.001 | 1.52 (1.19) |
| All encounters | 1.03 (1.26) | 0.931 (1.18) | <0.001 | 0.951 (1.20) |
| **Mean visits per quarter by type (2018-2019)** |  |  |  |  |
| Inpatient (include emergency department) | 0.209 (0.579) | 0.164 (0.526) | <0.001 | 0.173 (0.537) |
| Outpatient | 0.960 (1.20) | 0.871 (1.13) | <0.001 | 0.888 (1.15) |
| **Follow-up duration, years since Jan. 2020 Mean (SD)** | 3.01 (1.44) | 2.42 (1.65) | <0.001 | 2.53 (1.63) |
| **Total patient-quarters Mean (SD)** | 12.8 (9.50) | 11.6 (9.83) | <0.001 | 11.8 (9.77) |

\*Age is calculated using the age at the first visits.

Healthcare utilization patterns before the pandemic were similar. LC patients had higher baseline healthcare spending, both in mean quarterly cost ($886 vs. $775) and median quarterly cost ($232 vs. $154), and incurred slightly more total encounters per quarter (1.03 vs. 0.93). Differences were observed across service categories: LC patients had more inpatient or emergency visits (0.209 vs. 0.164 per quarter) and more outpatient encounters (0.960 vs. 0.871 per quarter). Hospitalizations during the baseline period were more frequent among LC patients (10.5% vs. 8.3%). LC patients contributed longer follow-up time after January 2020 (mean 3.01 vs. 2.42 years), with correspondingly more patient-quarters of data (12.8 vs. 11.6).

### Primary analysis: 2-part GEE censored at death

In the logistic component of the two-part model (probability of any quarterly healthcare use), Long COVID was associated with a 20% higher odds of having any healthcare utilization compared with individuals without PASC (OR 1.201, 95% CI: (1.175,1.227), p < 0.001). This difference increased over time, as shown by the significant PASC and quarter interaction (OR 1.014, 95% CI: (1.012,1.015), p < 0.01), indicating a progressive widening of utilization gaps across the post-acute period. Severity was also associated with healthcare utilization: patients hospitalized had 21% higher odds of quarterly healthcare utilization (OR 1.211, 95% CI: (1.183, 1.240), p < 0.0101). Additional factors associated with increased utilization included higher comorbidity burden, prior baseline spending, and non-Hispanic ethnicity, while male sex was associated with lower odds of use.

In the cost component, PASC was associated with 29.5% higher quarterly costs among patient-quarters with nonzero spending (Cost Ratio (CR) 1.295, 95% CI: (1.246, 1.346), p < 0.001). Hospitalization was associated with a 55% increase in conditional costs (CR 1.552, 95% CI: (1.507,1.598), p < 0.001). Male sex (CR 1.186), comorbidity burden (CR 1.106), and higher pre-pandemic spending (CR 1.203) were also associated with greater cost intensity. The PASC and quarter interaction (CR 1.005, 95% CI, (1.001, 1.008), p = 0.007) showed statistically significantly larger differences in the cost gap over time.

Adjusted predicted mean quarterly healthcare expenditures increased over time in both groups, with consistently higher costs among individuals with Long COVID (Figure 1). At baseline (Quarter 0), mean predicted expenditures were approximately $244 (95% CI, $224 to $264) for individuals with Long COVID and $164 (95% CI, $146 to $177) for those without Long COVID. By Quarter 19, these values rose to approximately $461 (95% CI, $422 to $498) and $225 (95% CI, $211 to $245), respectively.

**Figure 1.**
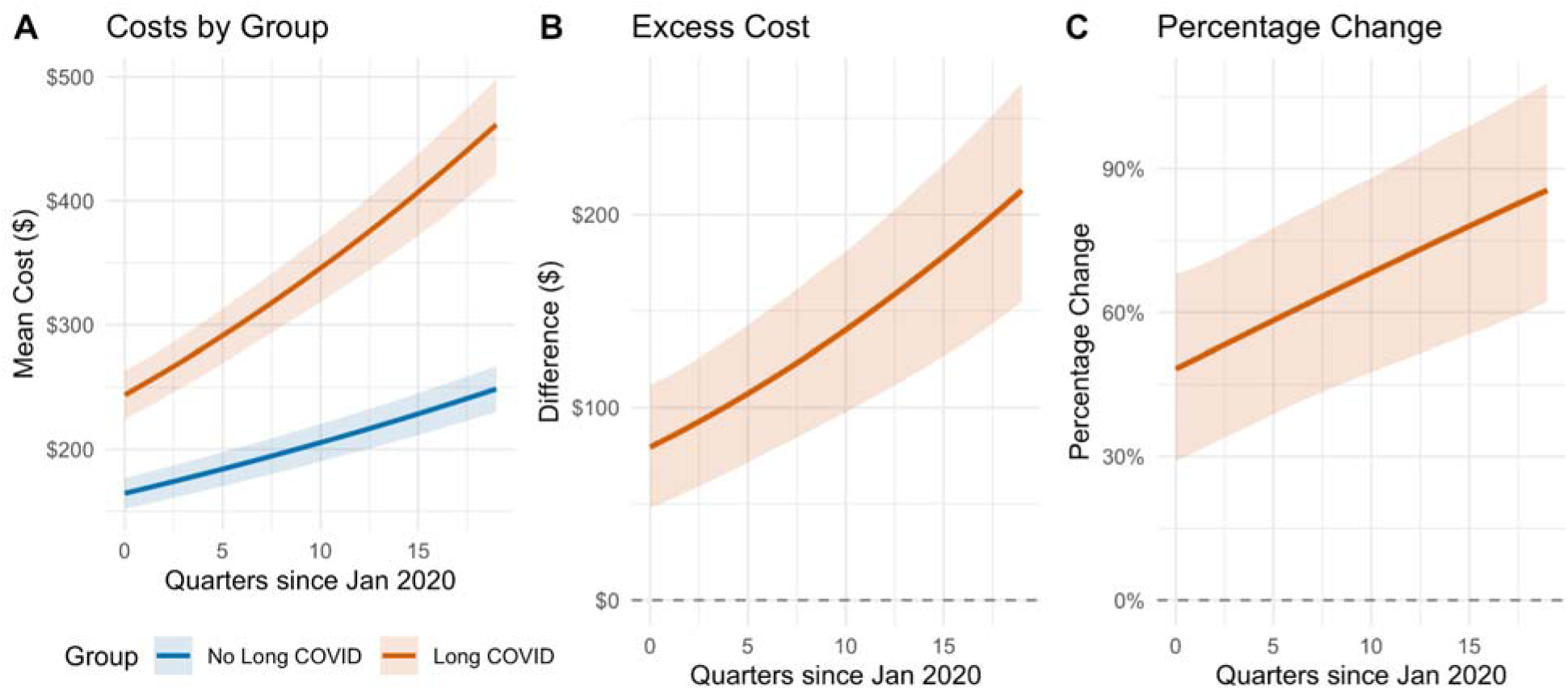
Adjusted predicted mean quarterly costs.

The adjusted cost difference between groups also widened steadily over the follow-up period. At baseline, individuals with Long COVID incurred an additional $79 per quarter (95% CI, $48 to $118) compared with those without Long COVID. By Quarter 19, this excess reached $236 per quarter (95% CI, $176 to $287). At the midpoint of follow-up (Quarter 10), the excess cost was $78 per quarter (95% CI, $31 to $133). The cost trajectory showed no evidence of plateauing; the rate of divergence between groups remained positive through the final observation period.

### Cost distribution

Healthcare costs exhibited substantial heterogeneity across patients, with excess costs concentrated in the upper tail of the distribution (Table 3). Among patient-quarters with any healthcare utilization, the cost distribution revealed marked heterogeneity in Long COVID’s economic impact. At the 25th percentile, costs were nearly identical between groups ($116 for Long COVID vs. $88 for controls, 32.2% difference). At the median, patients with Long COVID incurred $381, compared to $238 for controls, representing a $143 difference, or 60% higher costs. However, the excess burden increased substantially at higher percentiles: at the 90th percentile, the difference was $1,649 ($3,803 vs. $2,154, 76.6% higher), and at the 99th percentile, the difference reached $8,482 ($21,321 vs. $12,839, 66.1% higher).

**Table 3.** Distribution of Quarterly Healthcare Costs for any healthcare utilization.

| Percentile | Long COVID | No Long COVID | Difference | % Change |
| --- | --- | --- | --- | --- |
| 25th | \$ 116 | \$ 88 | \$ 28 | 32.2% |
| 50th (Median) | \$ 381 | \$ 238 | \$ 143 | 60.0% |
| 75th | \$ 1,224 | \$ 754 | \$ 470 | 62.3% |
| 90th | \$ 3,803 | \$ 2,154 | \$1,649 | 76.6% |
| 95th | \$ 7,451 | \$ 4,412 | \$3,038 | 68.9% |
| 99th | \$21,321 | \$12,839 | \$8,482 | 66.1% |
| **Mean** | **\$ 1,679** | **\$ 988** | **\$ 691** | **69.9%** |

### Visits Frequency

To assess the effect of increasing costs, we decomposed the overall cost trajectory into healthcare utilization (number of visits) and care intensity (cost per visit) components (Figure 2).

**Figure 2.**
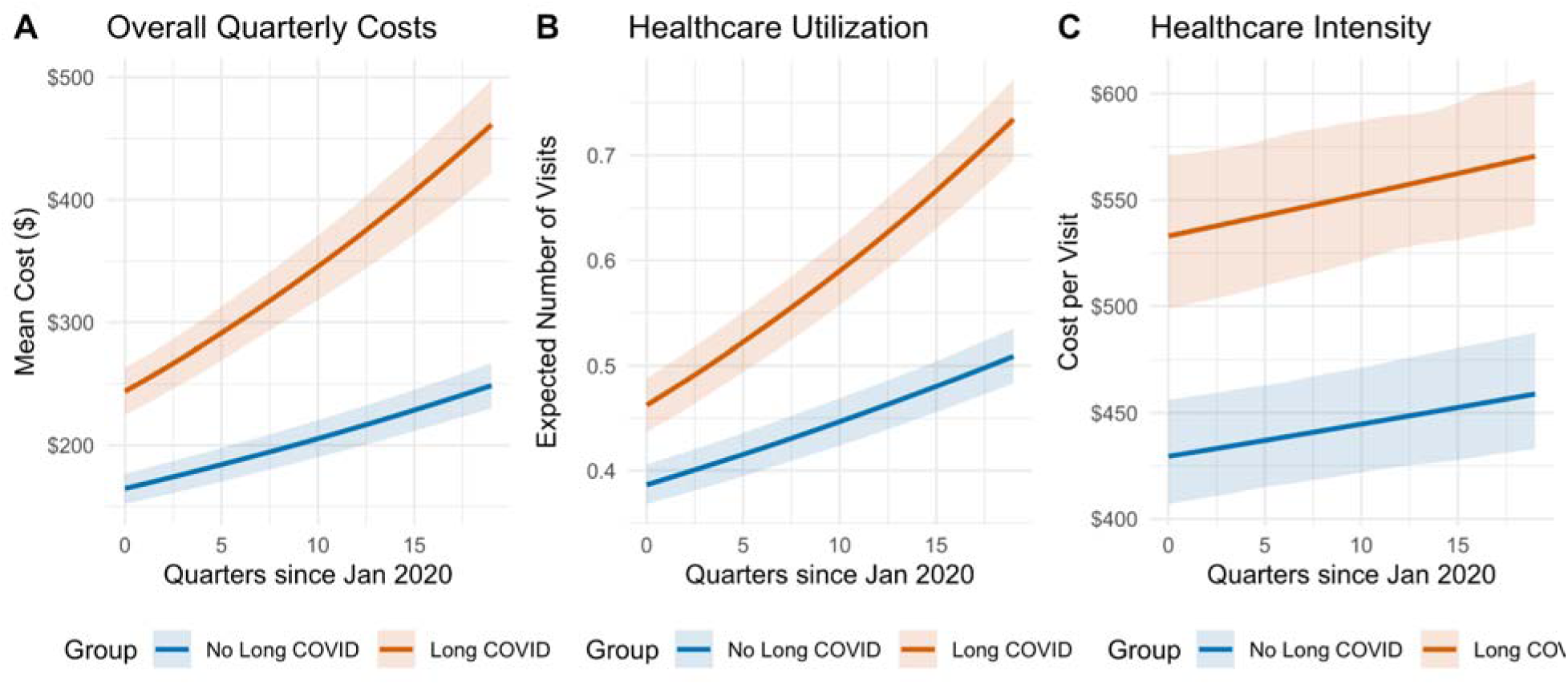
Predicted visit frequency and cost by long COVID status.

Patients with Long COVID demonstrated substantially higher healthcare utilization throughout the follow-up period (Figure 2B). At Quarter 0, Long COVID patients averaged 0.46 visits per quarter (95% CI, 0.44–0.49) compared with 0.39 visits (95% CI, 0.37–0.41) in those without Long COVID. By Quarter 19, these values increased to 0.73 visits (95% CI, 0.70–0.77) versus 0.51 visits (95% CI, 0.48–0.54), a 44% relative increase in visit frequency. The Poisson GEE model showed significant main effects for Long COVID status (RR: 1.26, 95% CI: 1.24-1.28, p<0.001) and a positive interaction with time (RR per quarter: 1.012, 95% CI: 1.011-1.013, p<0.001), indicating accelerating larger gaps in healthcare utilization over time.

Care intensity was consistently higher for Long COVID patients, measured as cost per visit among quarters with any healthcare use (Figure 2C). At baseline, cost per visit was $537 (95% CI: $523-$551) for Long COVID patients versus $368 (95% CI: $360-$376) for controls, representing a 46% premium. By quarter 19, these values were $569 (95% CI: $549-$590) versus $447 (95% CI: $438-$456), a 27% difference. The Gamma GEE model for cost per visit demonstrated significant Long COVID effects (cost ratio [CR]: 1.41, 95% CI: 1.38-1.44, p<0.001), though the interaction with time was small (CR per quarter: 0.998, 95% CI: 0.997-0.999, p=0.002), suggesting that while Long COVID visits remained more expensive throughout follow-up, the intensity premium remained relatively stable over time.

### Subgroup analysis

Long COVID was associated with persistently higher quarterly healthcare costs that widened over time. The widening cost trajectory was consistent across key demographic and clinical subgroups (Figure 3). Among patients stratified by infection severity, both non-hospitalized and hospitalized patients exhibited gradual increases in excess costs over time (Figure 3A). For non-hospitalized patients, excess costs increased from $62 (95% CI: $39–$85) at quarter 0 to $172 (95% CI: $137–$207) at quarter 19.

**Figure 3.**
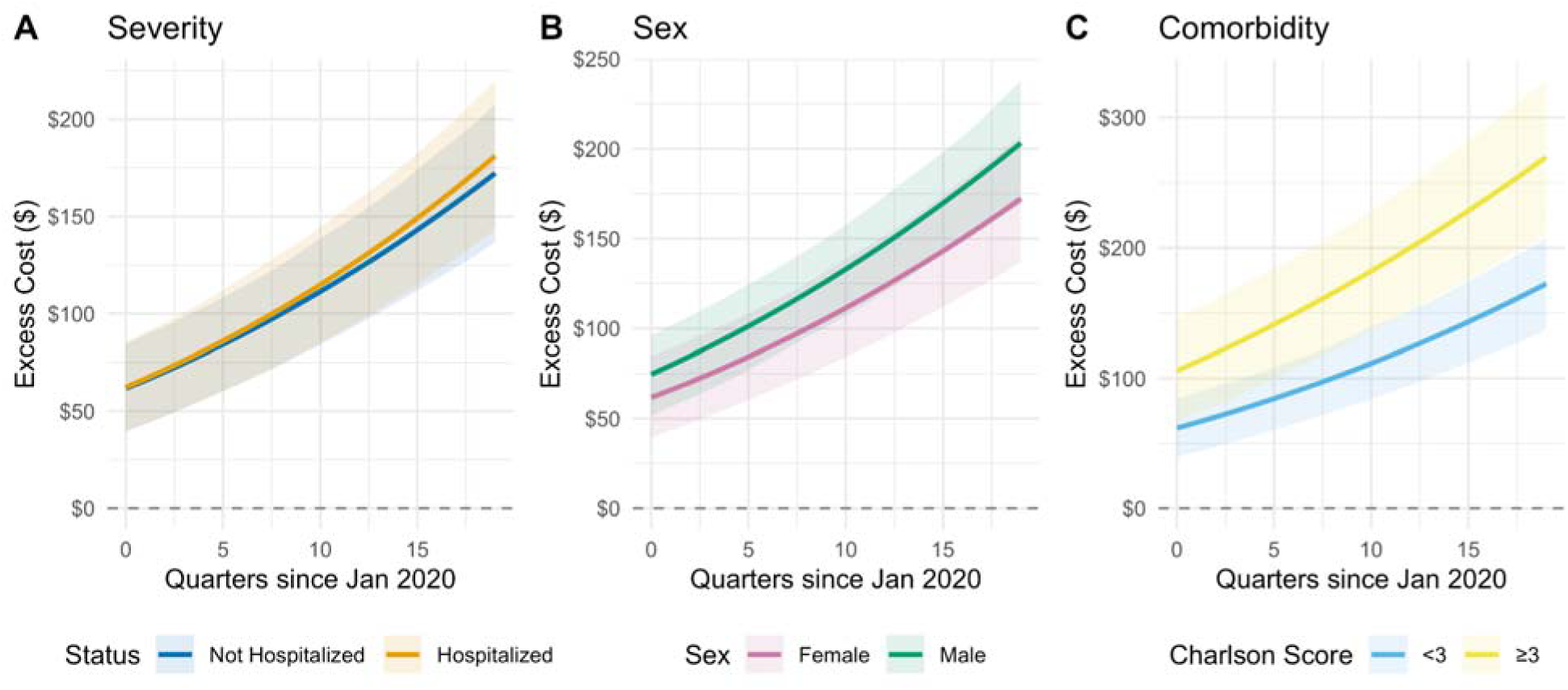
Cost Trajectories by disease severity, sex, and Comorbidity Burden.

Hospitalized patients, representing more severe acute COVID-19, showed higher absolute excess costs throughout follow-up, increasing from $62 (95% CI: $40–$86) to $181 (95% CI: $143–$220). Importantly, both groups demonstrated similar temporal slopes, indicating that the widening gap was not solely attributable to patients with severe initial infection.

Sex-stratified analyses revealed parallel cost trajectories for males and females (Figure 3B). Female Long COVID patients had excess costs of $62 (95% CI: $39–$85) at baseline, increasing to $173 (95% CI: $137–$209) by quarter 19. Male patients showed comparable patterns, with excess costs rising from $75 (95% CI: $51–$96) to $203 (95% CI: $169–$239).

Stratification by comorbidity burden further demonstrated consistency of the cost trajectory (Figure 3C). Patients with lower comorbidity burden (Charlson score <3) experienced increasing excess costs from $62 (95% CI: $39–$85) at quarter 0 to $173 (95% CI: $137–$209) at quarter 19. Those with higher comorbidity burden (Charlson score ≥3) had substantially higher excess costs throughout, increasing from $106 (95% CI: $67–$147) to $270 (95% CI: $228–$313). While absolute differences varied by comorbidity level, both groups exhibited the characteristic widening pattern over time.

### Sensitivity analysis

At baseline (Quarter 0), population-level expected costs were $543 for Long COVID patients and $407 for those without Long COVID (Figure 4). The Long COVID group exhibited a peak around Quarter 12 at approximately $1,037 (95% CI roughly $900–$1,150), while the NoLC group’s costs remained more stable, ranging from $430–$600 throughout follow-up. The declining survival probability over time was more pronounced in the Long COVID group, contributing to widening population-level cost differences.

**Figure 4.**
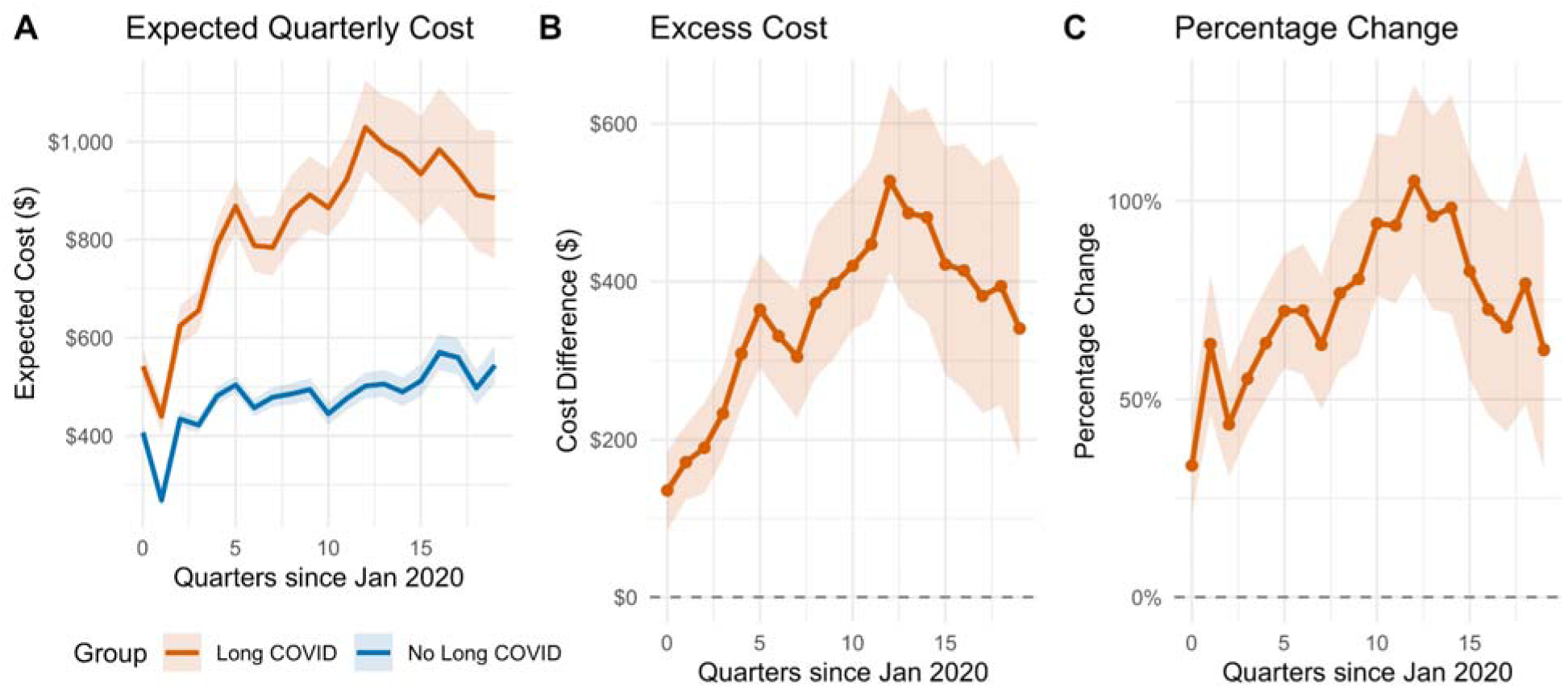
Population-level expected quarterly costs adjusted by mortality.

To account for mortality between groups, we estimated population-level healthcare costs and observed that patients with Long COVID incurred higher quarterly healthcare costs than patients without Long COVID over time. Over five years, the cumulative expected healthcare costs were $16,663 per Long COVID patient, about $7,124 higher than patients without Long COVID. Quarterly excess spending for Long COVID patients began shortly after infection and increased steadily over the first three years, peaking near Quarter 12 at approximately $397 (95% CI roughly $303–$501) of additional cost per patient, before gradually declining but remaining consistently positive through Year 5.

The decline in population-level costs after Quarter 12 reflects the combined effects of decreasing survival probability over time, potential improvements in healthcare utilization among survivors, and a changing composition of the surviving cohort. Importantly, the excess costs attributable to Long COVID remained substantial throughout follow-up, ranging from $340-480 per quarter in Years 4-5 (quarters 16-19), representing a persistent 63-79% cost premium relative to patients without Long COVID.

## Discussion

In this large retrospective cohort study of adults with documented SARS-CoV-2 infection, we found that Long COVID was associated with progressively widening healthcare costs over five years of follow-up. The adjusted excess quarterly cost increased from $79 at baseline to $236 by quarter 19, representing a threefold expansion of the cost gap. This trajectory was primarily driven by increasing healthcare utilization, rather than greater care intensity per encounter, and the pattern was consistent across subgroups defined by initial disease severity, sex, and comorbidity burden. After accounting for differential mortality, cumulative excess costs reached $7,124 per Long COVID patient over the five-year observation period.

These findings contradict the prevailing assumption that the economic burden of Long COVID represents a transient post-acute state that attenuates over time. Prior studies with follow-up limited to 6–15 months have documented elevated but relatively stable excess costs, ranging from 5% to 13% increases in difference-in-differences analyses. Our observation of progressively widening costs over five years, with the cost differential tripling from $79 to $236 per quarter, suggests that Long COVID behaves as a chronic, progressive condition with accumulating healthcare needs, rather than a post-acute state with resolving symptoms. The trajectory showed no inflection point suggesting stabilization, even at 60 months of follow-up. This distinction carries important implications for health system planning, as static cost projections based on short-term estimates will substantially underestimate the true economic burden.

The decomposition of costs into utilization and intensity components provides mechanistic insight into the nature of Long COVID’s economic impact. The widening gap in visit frequency, reaching 44% higher utilization among Long COVID patients by quarter 19, indicates an expanding need for ongoing clinical engagement. In contrast, the relatively stable cost-per-visit premium suggests that individual encounters are not becoming progressively more complex or resource-intensive. This pattern is consistent with chronic disease management characterized by frequent monitoring, symptom management, and care coordination rather than episodic acute exacerbations requiring intensive intervention. The shift from inpatient to outpatient cost drivers observed in prior studies aligns with this interpretation.

Several alternative explanations for the widening trajectory warrant consideration. First, differential mortality could create survivor bias if sicker Long COVID patients die earlier, but our mortality-adjusted sensitivity analysis demonstrated that the widening pattern persisted after accounting for survival differences. Second, the pattern could reflect healthcare-seeking behavior rather than true clinical need, but our adjustment for pre-pandemic utilization and the consistency across hospitalization subgroups argue against this explanation. Third, system-level factors, such as post-pandemic capacity recovery, could inflate later costs; however, both groups would be affected similarly, thereby preserving the between-group comparison. The most parsimonious interpretation is that Long COVID produces accumulating clinical complications or progressive functional decline requiring escalating healthcare engagement.

The concentration of excess costs in the upper tail of the distribution has implications for risk adjustment and care management strategies. At the 99th percentile, the absolute difference between groups exceeded $8,000 per quarter, indicating that a subset of Long COVID patients incur substantial and persistent high-cost care. Identifying characteristics that predict membership in this high-cost subgroup could inform targeted interventions and resource allocation. The consistency of the widening trajectory across hospitalization status subgroups is notable: patients who were not hospitalized during acute infection, representing the vast majority of SARS-CoV-2 infections, demonstrated cost patterns similar to those with severe initial disease. This finding underscores that the economic burden of Long COVID is not confined to patients with severe acute illness, and that the population-level impact extends across the full spectrum of initial disease severity.

Our estimates of excess costs fall between the lower bounds reported in French primary care data (€163 per year) and the upper bounds from economic modeling based on chronic fatigue syndrome parallels ($9,000 per year). This intermediate position likely reflects both methodological factors and true population differences. The precision phenotyping algorithm used to identify Long COVID in our cohort, which required symptom emergence following infection without explanatory prior diagnoses, may provide greater specificity than diagnosis code–based definitions while capturing a broader population than symptom survey approaches. The integrated health system setting, with comprehensive longitudinal records, enabled the complete capture of healthcare encounters across inpatient and outpatient settings. However, as discussed below, important cost components were not captured.

The declining trend in population-level costs after Year 3 likely reflects a combination of true clinical improvement in some patients, survivorship effects (with higher-cost patients experiencing earlier mortality), and the decreasing size of the at-risk cohort over time. The persistent excess costs throughout the entire five-year follow-up period demonstrate that the economic burden remains substantial for survivors, with Long COVID patients incurring 63-79% higher quarterly costs, even in Years 4 and 5. This pattern suggests that while some patients may experience symptom resolution or stabilization, Long COVID imposes a durable cost premium that extends well beyond the acute and post-acute phases of infection.

The findings have implications for clinical care, health policy, and research priorities. Clinically, the progressive nature of cost accumulation suggests that Long COVID patients may benefit from proactive longitudinal management strategies rather than reactive episodic care. Structured follow-up programs, multidisciplinary clinics, and care coordination interventions warrant evaluation for their potential to improve outcomes and reduce downstream costs. From a policy perspective, the magnitude and trajectory of excess costs, exceeding $7,000 per patient over five years among survivors, underscore the need for sustained investment in Long COVID research, clinical infrastructure, and disability support systems. Current estimates suggest that Long COVID affects tens of millions of individuals in the United States alone, implying aggregate costs measured in tens of billions of dollars annually, with implications for insurance risk pools, employer health expenditures, and public program budgets. For research, the widening cost trajectory underscores the importance of extended follow-up in Long COVID studies and raises questions about the underlying biological mechanisms driving the progressive clinical and economic burden.

### Limitations

This study has several limitations. First, healthcare costs were derived from procedure-based billing records and do not include medications, laboratory tests, or evaluation and management visit fees. This approach captures procedural and facility costs but underestimates total healthcare expenditures, particularly for conditions managed primarily through pharmacotherapy or frequent low-intensity visits. The true excess cost burden of Long COVID is therefore likely higher than our estimates, and the relative contribution of different cost components remains uncharacterized. Second, the study population was drawn from a single integrated health system in Massachusetts, which may limit the generalizability of the findings to other geographic regions, healthcare settings, and populations with different demographic compositions or insurance coverage patterns. Third, although we employed a validated precision phenotyping algorithm for the identification of Long COVID, misclassification remains possible in both directions. Some patients with Long COVID may not have been captured if their symptoms were attributed to pre-existing conditions, while some controls may have had unrecognized Long COVID. Such misclassification would generally bias estimates toward the null. Fourth, we were unable to capture healthcare utilization occurring outside the Mass General Brigham system, which could differentially affect cost estimates if patients with Long COVID sought care at external facilities at a rate different from that of controls. Fifth, the observational design precludes causal inference, and residual confounding by unmeasured factors associated with both the development of Long COVID and healthcare costs cannot be excluded. Sixth, our analysis did not capture indirect costs, including lost productivity, reduced work hours, and informal caregiving burden, which prior estimates suggest may exceed direct medical costs by a factor of two or more. Finally, the cohort includes patients infected across multiple pandemic waves with different viral variants and varying vaccination statuses; we did not stratify by these factors. The cost trajectory may differ for infections occurring in later periods, characterized by different viral characteristics and higher population immunity.

### Conclusions

In this large cohort study with a five-year follow-up, Long COVID was associated with healthcare costs that increased progressively over time, driven primarily by rising utilization rather than greater care intensity. The excess cost burden was consistent across subgroups and concentrated among high-cost patients. These findings indicate that Long COVID imposes a substantial and growing economic burden on the healthcare system, with implications for clinical management, health policy, and resource allocation. Extended follow-up studies and health economic evaluations incorporating the full spectrum of direct and indirect costs are needed to fully characterize the societal impact of this condition.

## Data Availability

Protected Health Information (PHI) restrictions limit the availability of the clinical data, which were used under IRB approval (protocol 2020P001063) for the current study only. As a result, this dataset is not publicly available.

## Ethics approval

Use of patient data in this study was approved by the Mass General Brigham Institutional Review Board (protocol 2020P001063).

## Acknowledgments

This study has been supported by grants from the National Institute of Allergy and Infectious Diseases (NIAID) R01AI165535.

