## Supplemental Table 1 for "Progressively Widening Healthcare Costs in Long COVID Over Five Years"

**Table 1S. Estimated effects from two-part GEE model (Long COVID vs No Long COVID)**

| Model | Term | Estimate | 95% CI | p-value |
| --- | --- | --- | --- | --- |
| Part 1: Any healthcare utilization (Estimate: Odds Ratio) | PASC vs No PASC | 1.201 | (1.175,1.227) | <0.001 |
|  | Quarter since Jan 2020 | 1.022 | (1.022,1.023) | <0.001 |
|  | Black vs. White | 0.987 | (0.965,1.010) | 0.275 |
|  | Asian vs. White | 0.879 | (0.849,0.910) | <0.001 |
|  | Other vs. White | 1.022 | (1.000,1.044) | 0.050 |
|  | Non-Hispanic vs. Hispanic | 1.079 | (1.046,1.112) | <0.001 |
|  | Hospitalized vs. Not hospitalized | 1.211 | (1.183,1.240) | <0.001 |
|  | Age | 0.992 | (0.991,0.993) | <0.001 |
|  | Age² | 1.000 | (1.000,1.000) | <0.001 |
|  | Male vs Female | 0.801 | (0.790,0.812) | <0.001 |
|  | Charlson | 1.195 | (1.191,1.200) | <0.001 |
|  | Log baseline quarterly cost | 1.107 | (1.101,1.112) | <0.001 |
|  | LC × Quarter interaction | 1.014 | (1.012,1.015) | <0.001 |
| Part 2: Cost \| use (Estimate: Cost Ratio) | PASC vs No PASC | 1.295 | (1.246,1.346) | <0.001 |
|  | Quarter since Jan 2020 | 1.006 | (1.004,1.008) | <0.001 |
|  | Black vs. White | 1.006 | (0.975,1.038) | 0.724 |
|  | Asian vs. White | 0.903 | (0.857,0.951) | <0.001 |
|  | Other vs. White | 0.940 | (0.910,0.970) | <0.001 |
|  | Non-Hispanic vs. Hispanic | 0.997 | (0.956,1.040) | 0.894 |
|  | Hospitalized vs. Not hospitalized | 1.552 | (1.507,1.598) | <0.001 |
|  | Age | 0.993 | (0.992,0.994) | <0.001 |
|  | Age² | 1.000 | (1.000,1.000) | <0.001 |
|  | Male vs Female | 1.186 | (1.163,1.209) | <0.001 |
|  | Charlson | 1.106 | (1.101,1.111) | <0.001 |
|  | Log baseline quarterly cost | 1.203 | (1.192,1.214) | <0.001 |
|  | LC × Quarter interaction | 1.005 | (1.001,1.008) | 0.007 |
